# Comparative Incidence and Outcomes of COVID-19 in Kidney or Kidney-Pancreas Transplant Recipients Versus Kidney or Kidney-Pancreas Waitlisted Patients: A Pilot Study

**DOI:** 10.1101/2020.07.20.20157990

**Authors:** Carlos A. Q. Santos, Yoona Rhee, Edward F. Hollinger, Oyedolamu K. Olaitan, Erik Schadde, Vasil Peev, Samuel N. Saltzberg, Martin Hertl

## Abstract

Comparative COVID-19 epidemiologic studies between immunosuppressed and immunocompetent patients may provide insight into the impact of immunosuppressive medications on clinical outcomes. In this retrospective cohort pilot study, we determined the incidence and outcomes of COVID-19 in kidney or kidney-pancreas transplant recipients and kidney or kidney-pancreas waitlisted patients in our center. COVID-19 testing was performed in 63 of 537 kidney or kidney-pancreas transplanted patients, versus 43 of 383 kidney or kidney-pancreas waitlisted patients as of June 4, 2020 (12% versus 11%, p=0.81). COVID-19 was identified in 14 of 537 kidney or kidney-pancreas transplanted patients, versus 9 of 383 kidney or kidney-pancreas waitlisted patients (2.6% versus 2.3%, p=0.81). Hospitalization occurred in 11 of 14 transplanted patients, and 4 of 9 waitlisted patients with COVID-19 (79% versus 44%, p=0.18). Intensive care unit admission occurred in 5 of 14 transplanted patients, and 1 of 9 waitlisted patients with COVID-19 (36% versus 11%, p=0.34). Two transplanted patients with COVID-19 were mechanically ventilated and died, whereas no waitlisted patients with COVID-19 died or were mechanically ventilated. Our study provides preliminary data that can be used for power calculations to inform multicenter studies designed to validate these findings.

## INTRODUCTION

Severe acute respiratory syndrome coronavirus 2 (SARS-CoV-2), the causative agent of coronavirus disease 2019 (COVID-19), poses significant threats to kidney transplant recipients. Case series and early cohort studies indicate an unfavorable disease course, with intensive care unit stays in 20% to 50% of patients, and death rates of 14% to 30%.^1-5^ These outcomes are worse than those reported for the general population, and may be due to greater age, immunosuppression, and comorbid conditions such as hypertension, diabetes mellitus and cardiovascular disease.^6^

The impact of chronic immunosuppression on COVID-19 outcomes has been debated. It has been hypothesized that a tempered immune response may prevent severe cytokine storm that ensues in a subset of patients with COVID-19 induced acute respiratory distress syndrome^.7,8^ However, chronic immunosuppression is known to worsen the disease courses of most infections, including but not limited to cytomegalovirus, influenza, *Streptococcus pneumoniae*, and invasive fungal infections.^9-12^ Epidemiologic studies examining disease courses of COVID-19 between comparable immunosuppressed and immunocompetent patients may provide insight regarding the impact of chronic immunosuppressive therapy on clinical outcomes.

In this retrospective cohort pilot study, we determined the incidence and outcomes of COVID-19 in kidney or kidney-pancreas transplant recipients and kidney or kidney-pancreas waitlisted patients at Rush University Medical Center in Chicago. We hypothesize that these patients have an incidence proportion of COVID-19 similar to the general population, but that transplanted patients have greater occurrences of hospitalization, intensive care unit admission, mechanical ventilation and death compared to waitlisted patients due to chronic immunosuppression. Understanding the comparative epidemiology of these otherwise matched populations may lead to insights regarding waitlist and transplant management during this pandemic.

## METHODS

### Study design and patient population

We performed a retrospective cohort study of kidney and kidney-pancreas transplant recipients who underwent transplantation from January 1, 2015 to April 30, 2020 (total n=560), and kidney and kidney-pancreas waitlisted patients from November 1, 2019 to April 30, 2020 (total n=476). We excluded transplant recipients who died before November 1, 2019 (n=23), since they could not have had the opportunity to develop COVID-19. We excluded waitlisted patients with a previous transplant since they had already been exposed to immunosuppression (n=93). The final study populations consisted of 537 transplant recipients, of whom 498 were kidney transplant recipients (372 deceased donor, 126 living donor) and 39 were kidney-pancreas transplant recipients, and 383 waitlisted patients, of whom 367 were waitlisted for kidney transplant and 16 were waitlisted for kidney-pancreas transplant. The study protocol was approved by the Institutional Review Board of Rush University Medical Center.

### Data sources

To facilitate obtaining data from the electronic health record in our center, we leveraged existing informatics infrastructure developed for the Chicago-Area Patient Centered Outcomes Research Network (CAPriCORN),^13^ a clinical data research network that is part of the Patient Centered Outcomes Research Network (PCORnet), and adapted it to populate normalized datasets with a daily refresh from November 1, 2019 to capture conditions and events during the COVID-19 pandemic.^14^ Demographics, healthcare encounters, vital signs, laboratory results, medications administered in the inpatient setting, and death were electronically collected according to CAPriCORN common data model specifications, and augmented with ancillary datasets on bed information to capture patient movement within hospital stays, and mechanical ventilation, to capture intubation and duration of ventilator use. Information outside of these domains were gathered by manual chart review of free text notes in the electronic health record using a standardized data collection tool by one physician-epidemiologist author (YR), and validated for accuracy by another physician-epidemiologist author (CS).

### COVID-19 identification, clinical characteristics and outcomes

COVID-19 was identified by querying the electronic datasets for detection of SARS-CoV-2 RNA by RT-PCR or nucleic acid amplification from nasopharyngeal swab specimens from November 1, 2019 to June 4, 2020. Demographic data, vital signs (temperature and body mass index), laboratory results, hospitalization, intensive care unit admission, mechanical ventilation, and death were captured from the electronic datasets, whereas comorbidities, organ type, cause of renal disease, induction and maintenance immunosuppression, presenting symptoms for COVID-19, oxygen supplementation, chest radiographic findings, and treatments tried for COVID-19 were collected by manual chart review. Demographics, comorbidities and maintenance immunosuppression were determined at the time of COVID-19 identification, whereas cause of renal disease and induction immunosuppression were determined at the time of transplant. All laboratory tests, radiologic evaluations and treatments were performed at the discretion of the healthcare team. Laboratory values were obtained from one day prior to 30 days after COVID-19 identification. Outcomes such as hospitalization, intensive care unit admission, mechanical ventilation, and death were identified with at least a 30-day follow-up censored for death or July 4, 2020, the time for data cutoff. These outcomes were first identified electronically, and then validated with manual chart review to ensure accuracy.

### Statistical analysis

Descriptive statistics were used to describe the demographic and clinical characteristics of the study populations, the incidence proportions of COVID-19 in the study populations, as well as the clinical features and outcomes of COVID-19. Chi-square or Fisher’s exact test were used as appropriate to determine if there were nonrandom associations between categorical variables. All analyses were performed in SAS version 9.3 (Cary, North Carolina).

## RESULTS

### Incidence proportion of COVID-19

COVID-19 testing was performed in 63 of 537 kidney or kidney-pancreas transplanted patients, equaling a testing proportion of 12%, compared to 43 of 383 kidney or kidney-pancreas waitlisted patients, equaling a testing proportion of 11%, as of June 4, 2020 (p=0.81). COVID-19 was identified in 14 of 537 kidney or kidney-pancreas transplanted patients, equaling an incidence proportion of 2.6%, compared to 9 of 383 kidney or kidney-pancreas waitlisted patients, equaling an incidence proportion of 2.3% (p=0.81). The first and latest cases of transplanted patients with COVID-19 were identified on March 25, 2020 and June, 1, 2020 respectively, and the first and latest waitlisted patients with COVID-19 were identified on March, 28, 2020 and June 3, 2020 respectively. For reference, COVID-19 testing was performed in 212,618 of 2,705,988 persons in Chicago, Illinois, equaling a testing proportion of 7.9%, and it was identified in 40,939 persons, equaling an incidence proportion of 1.5%, as of June 4, 2020 (personal communication, Stephanie Black, MD, Chicago Department of Public Health).

### Demographics and baseline characteristics of persons with COVID-19

The median ages of transplanted and waitlisted patients with COVID-19 were 52 and 49 years, respectively (**Table 1**). Transplanted and waitlisted patients with COVID-19 were predominantly male (64% versus 78%), African-American (57% versus 56%), non-Hispanic (57% versus 67%), and kidney transplanted or waitlisted (86% versus 89%). Common causes of renal disease for transplanted and waitlisted patients were diabetic nephropathy (43% versus 78%) and hypertensive nephroangiosclerosis (29% versus 11%), and commonly identified comorbidities were hypertension (93% versus 100%), diabetes mellitus (50% versus 78%) and heart disease (57% versus 56%). The median body mass index (BMI) for transplanted and waitlisted patients with COVID-19 were 27.3 and 28.3 kg/m^2^ respectively. Alemtuzumab was more frequently used than anti-thymocyte globulin in transplanted patients for induction immunosuppression (57% versus 43%), and maintenance immunosuppression typically consisted of a calcineurin inhibitor (tacrolimus or cyclosporine), an antimetabolite (mycophenolate or azathioprine), and prednisone.

**Table 1.** Demographics and baseline characteristics of kidney or kidney-pancreas transplant recipients and kidney or kidney-pancreas waitlisted patients with COVID-19.

| **Variable \*** | **Kidney or Kidney-Pancreas Transplant Recipients** | **Kidney or Kidney-Pancreas Waitlisted Patients** |
| --- | --- | --- |
| **Age – median (range) †** | 52 (31 – 72) | 49 (36 – 73) |
| **Demographics – no./total no. (%)** |  |  |
| Male | 9/14 (64) | 7/9 (78) |
| African-American | 8/14 (57) | 5/9 (56) |
| Non-Hispanic | 8/14 (57) | 6/9 (67) |
| **Organ type – no./total no. (%)** |  |  |
| Kidney | 12/14 (86) | 8/9 (89) |
| Kidney-pancreas | 2/14 (14) | 1/9 (11) |
| **Cause of renal disease – no./total no. (%)** |  |  |
| Diabetic nephropathy | 6/14 (43) | 7/9 (78) |
| Glomerulonephritis | 2/14 (14) | 0/9 (0) |
| Hypertensive nephroangiosclerosis | 4/14 (29) | 1/9 (11) |
| Others | 2/14 (14) | 2/9 (22) |
| **Comorbidities – no./total no. (%)** |  |  |
| Hypertension | 13/14 (93) | 9/9 (100) |
| Diabetes mellitus | 7/14 (50) | 7/9 (78) |
| Heart disease | 8/14 (57) | 5/9 (56) |
| Lung disease | 1/14 (7) | 1/9 (11) |
| Cancer | 1/14 (7) | 0/9 (0) |
| Smoking | 1/14 (7) | 1/9 (11) |
| **BMI – median (range) – kg/m<sup>2</sup> ‡** | 27.3 (18.6 – 41.9) | 28.3 (20.6 – 34.2) |
| **Induction immunosuppression – no./total no. (%)** |  |  |
| Alemtuzumab | 8/14 (57) |  |
| Anti-thymocyte globulin | 6/14 (43) |  |
| Basiliximab | 0/14 (0) |  |
| High-dose corticosteroids | 14/14 (100) |  |
| **Maintenance immunosuppression – no./total no. (%)** |  |  |
| Tacrolimus or Cyclosporine | 13/14 (93) |  |
| Mycophenolate or Azathioprine | 12/14 (86) |  |
| Prednisone | 12/14 (86) |  |
| Sirolimus | 2/14 (14) |  |
| Everolimus | 0/14 (0) |  |

\* Age, demographics and BMI were derived from electronically generated datasets mapped to CAPriCORN common data model specifications; organ type, cause of renal disease, comorbidities, induction immunosuppression and maintenance immunosuppression were identified by manual chart review
† Age at time of COVID-19 identification
‡ Most recent BMI prior to COVID-19 identification
COVID-19 indicates coronavirus disease 2019; BMI, body mass index; CAPriCORN, Chicago Area Patient Centered Outcomes Research Network

### Clinical and laboratory features of COVID-19

Common presenting symptoms of COVID-19 in transplanted and waitlisted patients were fever (64% versus 67%) and cough (43% and 56%) (**Table 2**). Supplementary oxygen was provided to 36% of transplanted patients, and 33% of waitlisted patients. Very high grade fever greater than 104°F was identified in one kidney transplanted patient. Frequent laboratory abnormalities in transplanted and waitlisted patients with COVID-19 were leukopenia (38% versus 67%), lymphopenia (85% versus 50%), thrombocytopenia (31% versus 50%), and elevations in aspartate aminotransferase (33% versus 60%), C-reactive protein (86% versus 100%), ferritin (64% versus 100%) and D-dimer (100% versus 67%). Among transplanted patients on an antimetabolite for immunosuppression, the antimetabolite was withdrawn in 83% of patients. Transplanted and waitlisted patients were infrequently given hydroxychloroquine (14% versus 0%) or high-dose glucocorticoids (14% versus 0%). Tocilizumab was used in one kidney transplanted and one kidney waitlisted patient, and remdesivir was administered to one kidney transplanted patient.

**Table 2.** Clinical features and outcomes of COVID-19 in kidney or kidney-pancreas transplant recipients and kidney or kidney-pancreas waitlisted patients.

| **Variable \*** | **Kidney or Kidney-Pancreas Transplant Recipients** | **Kidney or Kidney-Pancreas Waitlisted Patients** |
| --- | --- | --- |
| **Presenting symptom – no./total no. (%)** |  |  |
| Fever | 9/14 (64) | 6/9 (67) |
| Cough | 6/14 (43) | 5/9 (56) |
| Dyspnea | 3/14 (21) | 2/9 (22) |
| Myalgias | 3/14 (21) | 2/9 (22) |
| Diarrhea | 2/14 (14) | 4/9 (44) |
| **Oxygen requirement – no./total no. (%)** | 5/14 (36) | 3/9 (33) |
| **Temperature > 104°F – no./total no. (%) †** | 1/14 (7) | 0/9 (0) |
| **Laboratory values †** |  |  |
| White-cell count |  |  |
| Median (range) – per mm<sup>3</sup> | 8,800 (2,610 – 30,910) | 4,540 (1,880 – 7,620) |
| Patients with count <4,000 per mm<sup>3</sup> (leukopenia) – no./total no. (%) | 5/13 (38) | 4/6 (67) |
| Lymphocyte count |  |  |
| Median (range) – per mm<sup>3</sup> | 740 (260 – 1,410) | 1,000 (420 – 1,800) |
| Patients with count <1000 per mm<sup>3</sup> (lymphopenia) – no./total no. (%) | 11/13 (85) | 3/6 (50) |
| Platelet count |  |  |
| Median (range) – per mm<sup>3</sup> | 223,000 (87,000 – 435,000) | 180,000 (63,000 – 448,000) |
| Patients with count <150,000 per mm<sup>3</sup> (thrombocytopenia) – no./total no. (%) | 4/13 (31) | 3/6 (50) |
| Aspartate aminotransferase |  |  |
| Median (range) – mg/dl | 20 (9 – 206) | 26 (12 – 62) |
| Patients with level > 50 mg/dl – no./total no. (%) | 4/12 (33) | 3/5 (60) |
| Alanine aminotransferase |  |  |
| Median (range) – mg/dl | 19 (6 – 70) | 14 (7 – 44) |
| Patients with level > 50 mg/dl – no./total no. (%) | 3/12 (25) | 0/5 (0) |
| C-reactive protein |  |  |
| Median (range) – mg/dl | 212.4 (10.2 – 396.2) | 162.1 (41.0 – 229.8) |
| Patients with level > 5 mg/dl – no./total no. (%) | 6/7 (86) | 3/3 (100) |
| Ferritin |  |  |
| Median (range) – mg/dl | 4,933 (298 – 25,677) | 1,544 (552 – 6,649) |
| Patients with level > 900 mg/dl – no./total no. (%) | 7/11 (64) | 3/3 (100) |
| D-dimer |  |  |
| Median (range) – mg/dl | 11.4 (5.3 – 57.8) | 7.4 (3.8 – 8.8) |
| Patients with level > 5 mg/dl – no./total no. (%) | 4/4 (100) | 2/3 (67) |
| **Chest radiographic findings consistent with viral pneumonia – no./total no. (%)** | 8/11 (73) | 4/5 (80) |
| **Treatment – no./total no. (%) \*\*** |  |  |
| Withdrawal of antimetabolite | 10/12 (83) |  |
| Withdrawal of calcineurin inhibitor | 3/13 (23) |  |
| Withdrawal of mTOR inhibitor | 2/2 (100) |  |
| Hydroxychloroquine | 2/14 (14) | 0/9 (0) |
| Tocilizumab | 1/14 (7) | 1/9 (11) |
| High-dose glucocorticoids | 2/14 (14) | 0/9 (0) |
| Remdesivir | 1/14 (7) | 0/9 (0) |
| **Outcomes at a median of 52 days of follow-up (range, 5-101) – no./total no. (%) ‡** |  |  |
| Hospitalization – no./total no. (%) | 11/14 (79) | 4/9 (44) |
| Intensive care unit admission – no./total no. (%) | 5/14 (36) | 1/9 (11) |
| Mechanical ventilation | 2/14 (14) | 0/9 (0) |
| Death | 2/14 (14) | 0/9 (0) |

\* Temperature, laboratory values and outcomes (hospitalization, intensive care unit admission, mechanical ventilation and death) were derived from electronically generated datasets mapped to CAPriCORN common data model specifications; outcomes were subsequently validated with manual chart review; presenting symptoms, oxygen requirement, chest radiographic findings and treatment were identified by manual chart review
† Body temperature and laboratory tests taken from one day prior through 30 days after COVID-19 identification; some patients did not have laboratory tests performed
\*\* Antimetabolite includes mycophenolate or azathioprine; calcineurin inhibitors include tacrolimus and cyclosporine; mTOR inhibitors include sirolimus and everolimus
‡ Hospitalization, intensive care unit admission and mechanical ventilation identified from three days prior through 30 days after COVID-19 identification
COVID-19 indicates coronavirus disease 2019; CAPriCORN, Chicago Area Patient Centered Outcomes Research Network

### Outcomes of COVID-19

Hospitalization occurred in 79% and 44% of transplanted and waitlisted patients with COVID-19 respectively (p=0.18). Intensive care unit admission occurred in 5 of 14 transplanted patients, and 1 of 9 waitlisted patients with COVID-19 (36% versus 11%, p=0.34). Two transplanted patients with COVID-19 were mechanically ventilated and died, whereas no waitlisted patients with COVID-19 died or were mechanically ventilated.

## DISCUSSION

We found in this retrospective cohort pilot study that the incidence proportion of diagnosed COVID-19 in kidney or kidney-pancreas transplanted patients was similar to that of kidney or kidney-pancreas waitlisted patients in our center as of June 4, 2020. We also found that transplanted patients had numerically more occurrences of hospitalization, intensive care unit admission, mechanical ventilation or death compared to waitlisted patients. Tests for nonrandom associations did not show statistical significance, but definitive conclusions will require adequately powered multicenter studies. The use of CAPriCORN and PCORnet common data models employed by this study to capture some data domains and outcomes promotes replicability in other centers, and may enable the conduct of adequately powered multicenter studies.

Previous epidemiologic studies on COVID-19 in kidney transplant recipients focused on assembling cohorts of patients diagnosed with COVID-19 and describing their clinical features and outcomes.^1-5^ These studies did not assemble cohorts of transplanted patients who were followed over time to identify COVID-19 infection, which precluded the determination of its incidence. Reports on COVID-19 in kidney waitlisted patients or patients with end-stage renal disease was limited to one case report and one case series of five patients on hemodialysis who contracted COVID-19.^15,16^ We found that the incidence proportions of COVID-19 in transplanted and waitlisted patients were comparable, and were slightly higher than that of the general population in our city. However, testing proportions for COVID-19 in transplanted and waitlisted patients were also higher than that of the general population, indicating either greater access to healthcare among transplanted or waitlisted patients relative to other persons in the community, or lower thresholds for testing among transplanted or waitlisted patients. Testing proportions for COVID-19 in transplanted and waitlisted patients were similar, indicating no assessment bias between cohorts.

The clinical features of COVID-19 in kidney transplanted and waitlisted patients were comparable, and similar to what has already been reported in the literature.^1-5,16^ Fever and cough were common presenting symptoms, but less common symptoms such as myalgias and diarrhea were also identified. Leukopenia, lymphopenia and thrombocytopenia were frequently found among patients in whom a complete blood count was done, and elevations in C-reactive protein and ferritin were prominent among patients in whom these markers of inflammation were checked. The most commonly tried treatment was discontinuation of the antimetabolite, similar to the management of viral infections such as cytomegalovirus or BK virus.^17,18^ Remdesivir was given to one kidney transplanted patient in this study as part of a randomized controlled trial who subsequently died. Preliminary results suggest that remdesivir administration is associated with shorter recovery times and a trend towards lower mortality.^19^ Further studies are needed to confirm these effects. Hydroxychloroquine was tried in two kidney transplant patients earlier in the course of the pandemic on account of *in vitro* activity against SARS-CoV-2.^20^ However, emerging evidence from observational studies and interim analyses of clinical trials evaluating the safety and effectiveness of hydroxychloroquine for COVID-19 treatment have shown either no or modestly deleterious effects,^21,22^ prompting the Federal Drug Administration to revoke emergency use authorization of this drug for COVID-19 treatment. Tocilizumab was tried in one kidney transplanted patient and one kidney waitlisted patient. The transplanted patient died, and the waitlisted patient survived. Tocilizumab is a human monoclonal IL-6 receptor antagonist used for treatment of rheumatoid arthritis, Castleman disease, and cytokine release syndrome due to chimeric antigen receptor T-cell therapy^.23,24^ The use of tocilizumab to modulate the exuberant and dysregulated inflammatory response among patients with COVID-19 associated acute respiratory distress syndrome has been reported in a case series of five solid-organ transplant recipients, of whom favorable short-term outcomes, including improvement in respiratory status, were achieved in four.^25^ Randomized controlled trials of tocilizumab for this indication are needed to determine its safety and effectiveness.

We found that transplanted patients had numerically more occurrences of hospitalization, intensive care unit admission, mechanical ventilation or death compared to waitlisted patients. Tests for nonrandom associations did not show statistical significance, but definitive conclusions will require adequately powered multicenter studies. The case fatality rates we found for transplanted and waitlisted patients with COVID-19 were 14% and 0% respectively, which is similar to reported rates in other studies of kidney transplant recipients,^1-5^ and patients on hemodialysis.^16^ Multicenter studies will be needed to validate the relative acquisition risks and case fatality rates of kidney transplanted and waitlisted patients that we found in this study. However, we provide preliminary data that can be used for power calculations to inform the design of multicenter studies. Presuming a 14% difference in mortality between kidney transplanted and waitlisted patients with COVID-19, and an alpha of 0.05 and beta of 0.2, 51 transplanted and 51 waitlisted patients with COVID-19 would need to be identified. If a more conservative estimate of 10% mortality difference is used, 73 transplanted and 73 waitlisted patients with COVID-19 would need to be identified. These numbers are likely achievable among the five transplant centers participating in CAPriCORN.^13^

Our preliminary findings may also inform clinical decision making on whether to transplant patients during the COVID-19 pandemic, or delay transplant until after the pandemic. A recent simulation and machine learning study identifying scenarios of benefit or harm from kidney transplantation during the COVID-19 pandemic showed that immediate kidney transplant provided survival benefit in most scenarios, and that kidney transplant only began showing evidence of harm in scenarios when case fatality rates for COVID-19 were greater than 50% among transplanted patients.^26^ The 14% case fatality rate we found in our center for kidney transplanted patients is below this threshold. If validated, it would indicate that kidney transplantation would benefit most patients during the COVID-19 pandemic if local resources allow.

The strengths of our study are direct comparison of kidney or kidney-pancreas transplanted and waitlisted patients with respect to incidence and outcomes of COVID-19, and the use of CAPriCORN and PCORnet common data models to capture these measures that promotes replicability in other centers. It however, has limitations. First, it is a single-center pilot study with low numbers, thereby making definitive conclusions difficult. However, it provides important preliminary data that can be used for power calculations to inform the design of multicenter studies. Second, the incidence proportions of COVID-19 identified for these patients represent minimum estimates since some cases of COVID-19 may have been identified outside of our healthcare system and missed by our electronic health record. However, minimum estimates are still useful as it provides the lowest possible incidence proportions for the populations of interest. Third, our findings may not be generalizable since it is a single-center study. However, it provides data that may inform the design of multicenter studies that may have more generalizable results.

In summary, we found that the incidence proportion of COVID-19 in kidney or kidney-pancreas transplanted patients was similar to that of kidney or kidney-pancreas waitlisted patients, and that transplanted patients had numerically more occurrences of hospitalization, intensive care unit admission, mechanical ventilation or death compared to waitlisted patients. Tests for nonrandom associations did not show statistical significance, but definitive conclusions will require adequately powered multicenter studies. This study provides preliminary data that can be used for power calculations for multicenter studies.

## Data Availability

The data that support the findings of this study are available on request from the corresponding author. The data are not publicly available due to privacy or ethical restrictions.

## ACKNOWLEDGMENTS

The authors would like to acknowledge Ronda Billerbeck, Ekta Kishen and Karthikeyan Swaminathan for their help with database management. No external funding was received.

## Abbreviations

BMI: body mass index
CAPriCORN: Chicago-Area Patient Centered Outcomes Research Network
COVID-19: coronavirus disease 2019
PCORnet: Patient Centered Outcomes Research Network
RNA: ribonucleic acid
RT-PCR: reverse transcription polymerase chain reaction
SARS-CoV-2: severe acute respiratory syndrome coronavirus 2

